# Compositional analysis of genetic ancestry shows that relative proportion of African ancestry is positively associated with fecal myeloperoxidase in Brazilian infants

**DOI:** 10.64898/2026.10.02.26364607

**Authors:** Josyf C. Mychaleckyj, Alexandre Havt, Uma Nayak, Richard L. Guerrant, Aldo A. M. Lima

**Affiliations:** Department of Genome Sciences, University of Virginia, Charlottesville, VA, USA; Departamento de Fisiologia e Farmacologia, Universidade Federal do Ceará, Fortaleza, Brazil; Center for Global Health, University of Virginia, Charlottesville, VA, USA; INCT-Instituto de Biomedicina, Universidade Federal do Ceará, Fortaleza, Brazil

**Keywords:** Genetic ancestry, admixture, genetics, diarrhea, compositional analysis, malnutrition, African ancestry, infectious disease, host genetics, Aitchison geometry

## Abstract

Genetic ancestry composition has been used extensively as a predictor of clinical outcomes, multiomic biomarkers, and endophenotypes, but the models have most often used untransformed ancestry fractions as explanatory variables. Under a simplex constraint results from this type of model can be uninterpretable and misleading. We demonstrate the use of multiple transformed log ratio coordinates of ancestral proportions as predictors in a study of excreted myeloperoxidase (MPO), a neutrophilic enzyme used as a fecal biomarker to measure intestinal inflammation and enteropathy. We show how to convert these difficult-to-interpret effects into association results for shifts between two ancestries under an isoancestry constraint (holding an ancestry fraction constant) using post-estimation average marginal effects and explain their geometric interpretation. We found that increasing proportion of African ancestry was associated with increased levels of excreted MPO in low-income household infants in three Northeastern Brazil studies, with a smaller negative secondary association of European ancestry, all under a tri-continental ancestry composition (African, European, Amerindian). While the maximum log(MPO) outcome variance explained by differences in genetic ancestry was modest at a few percent, we show evidence that these differences in heritability were context-dependent on measurement of MPO under active enteric infection. The results were attenuated by socio-economic status, but a signal remained. Overall, we found that African genomes carry additional alleles that enable a more intensive neutrophil response to enteric infection relative to European and Amerindian genomes, illuminating a possible host genetic background role in modulating immune response to enteric diseases and pathogens.

## Introduction

Myeloperoxidase (MPO) is a neutrophilic enzyme excreted in feces and linked to its critical biological role in protecting against microorganisms, there is considerable interest in the potential use of MPO as a non-invasive fecal biomarker to measure intestinal inflammation in environmental enteropathy (EE), inflammatory bowel disease, and other colitides ^1–6^. EE is a subclinical intestinal disorder characterized by mucosal and systemic inflammation, reduced intestinal barrier integrity and increased permeability, bacterial translocation, and reduced intestinal absorptive capacity ^7–9^. Infants in low and middle-income countries develop EE as a result of poor hygiene and sanitation, and hence repeated enteropathogen exposure ^5^; this puts the infants at risk for future impaired growth and cognitive development ^10, 11^.

The largest previous longitudinal cohort study of fecal MPO in infants was conducted by the ‘Etiology, Risk Factors, and Interactions of Enteric Infections and Malnutrition and the Consequences for Child Health and Development’ (MAL-ED) consortium, a global collaboration between laboratories including some of the studies and investigators of this work ^12^. They found multiple factors that were associated with higher fecal MPO levels, including age of the infant, breastfeeding, enteric pathogen load, and lower socio-economic status, with much smaller contributions from symptomatic clinical comorbidities and seasonality. However, the same study also noted that a large proportion of variance of MPO was not explained by the traditional factors considered. We hypothesized that genetic ancestry could be an additional factor explaining some of the variance in infant fecal MPO levels and sought to test our hypothesis using genetically-inferred ancestry in at-risk admixed Brazilian infants enrolled in our existing studies, recruited from favelas and low socio-economic households centered on Fortaleza, Ceará state, but also including surrounding states.

The genetic ancestry of an individual is completely specified as the ancestral lineages of inherited DNA through ancestral recombination graphs ^13^. In common genetic parlance these complexities are reduced to a simple concept of ancestry based on discrete continental (or subcontinental) population groupings which largely map to genetic compositional groups defined by genetic similarity^14–18^. While acknowledging the greater inheritance information and complexity of the former, the latter concept will be implied in this work. Testing association of genetic ancestry with traits has been useful to derive evidence for a genetic trait component; identifying and estimating the macroscopic genome-level contribution to population variation in a trait; disentangling the effects of race and genetics; and mapping the population genetic history of a trait-associated variant. Where statistical models have been used to infer these relationships, these have almost invariably used the discrete fractional genetic subgroup compositions as predictors.

It is well-known that compositional data such as genetic ancestry proportions do not conform to Euclidean geometry when used a basis set for a measure because the ever-present simplex constraint requires that proportions to sum to unity. Hence standard statistical models using untransformed proportions, even when entered into a model individually, can be misleading and even uninterpretable since it is impossible for a single component composition to vary independently ^19^. We applied rigorous statistical composition data analysis methods to investigate the relationship between genetic ancestry and excreted MPO levels in infants from Northeastern Brazil and develop what we believe is a new post-estimation analysis method to render the constrained log ratio results more interpretable. We discuss this in a global ancestry analysis but these analytic issues could be relevant in local ancestry analytical models also.

## Methods

### 2.1 Brazil Study Populations

The genetic samples analyzed in this work were drawn from 4 studies conducted on populations in Northeastern Brazil and centered on Fortaleza, Ceará state. They have been previously described in detail and will only be reviewed briefly here. The parent Malnutrition and Enteric Disease Network (Mal-ED) study is described in ^20^ while the longitudinal Mal-ED Birth Cohort (MBC) and prospective Mal-ED Case-Control (MCC) study groups were described elsewhere ^21^. The MBC study enrolled 292 children within 17 days of birth between 2010 to 2014 and they were followed up for 48 months. The MCC study enrolled 401 children from 6-18 months of age between 2010 to 2014 where cases were malnourished infants exhibiting a weight for age Z score (WAZ) of less than -2 and controls had WAZ higher than -1. The Recodisa (REC) Case-Control study was designed to analyze the etiology of diarrhea in the historically indebted semiarid region of Brazil and enrolled 1200 children from 2 to 36 months of age from 2010 to 2014 ^22^. They were all recruited from hospitals or clinic facilities sited in 6 semiarid countryside cities of more than 50,000 inhabitants in the Northeast of Brazil, including Crato (Ceará state); Cajazeiras, Souza and Patos (Paraíba state); Ouricuri (Pernambuco state); and Picos (Piauí state). Cases were ascertained as children with three or more episodes of diarrhea in the 24 hours preceding enrollment. Each city enrolled 100 cases and 100 controls from whom was collected one sample of stool and saliva and clinical anthropometric measurements and socioeconomic parameters.

### 2.2 Study Ethics

Study protocols were reviewed and approved by the Institutional Review Board for Health Sciences Research of the University of Virginia and the Research Ethics Committee of Universidade Federal de Ceará.

### 2.3 Stool Sample Collection, Fecal Myeloperoxidase Measurement, and Harmonization

The study groups were selected based on the availability and age of quantitative fecal MPO measurement. In the motivating MAL-ED longitudinal multi-cohort study of excreted fecal MPO in infants ^12^, the age-related mean MPO peaked around 3-6 months with a gradual decline thereafter to the study age limit of 24 months. This peaking likely represents gut maturation and closing in early infancy ^23, 24^, as well as a transition from maternal dependency for immune protection to increasing expression of the infant genetic factors. We chose a 12-24 month stool sample age to avoid these complicating developmental changes and to minimize age differences among studies. For the longitudinal MBC we included all MPO measures after one year of age. For MCC we selected the sample obtained within a one month window of infant participation start (approx. one year of age) where possible, else the first MPO available during follow up was taken. For the REC study, the single stool sample was obtained within a one month window of study start. The protocol and operational methods for the acquisition of stools and measurement of MPO within the Mal-ED birth cohorts have been described ^5^. Briefly, non-diarrheal episode surveillance stool samples were collected during the first year and quarterly in the second. All stool samples were collected without fixative by field workers and stored frozen at −70°C. The batched MPO assays used the vendor kit protocol (Alpco, Salem, NH) except initial dilutions were 1:500 in Brazil. Similar laboratory assay methods were used in the other studies with stools collected on different sampling schedules ^6, 22^. Finally, we dropped any MPO measurement from a diarrheal stool since the stools were invariably more liquid and standardization of the measured MPO quantity per stool weight is much less accurate.

### 2.4 Genome-wide Genotyping and Quality Control of Brazil DNA Samples

The genotyping and quality control of these samples has been described previously ^25^. We genotyped 2,119 samples on the Affymetrix Axiom LAT-1 array (818,154 SNPs) drawn from 3 studies in the Northeastern region of Brazil, centered on Fortaleza. Post-QC there were 1,538 samples and 755,801 SNPs annotated using Affymetrix release NA34 (hg19). These were joined to MPO phenotype and age, gender, and breastfeeding covariate data to realize the final analysis data sets.

### 2.5 Estimation of Genetic Ancestry

Herein, genetic ancestry refers to the estimated genome-wide composition of an individual genome based on partitioning the individual’s whole genome variation to discrete continental population groups through a process of comparing genetic similarity of mosaic genome segments. The methods for estimating the proportions of genetic ancestry in this study have been described previously ^25^. Unsupervised analysis on the combined genetic data set using ADMIXTURE 1.23 ^26, 27^ suggested a latent three ancestral cluster model best fitted the data, but since estimates from an unsupervised admixture analysis are known to be potentially biased and inaccurate ^28, 29^, we identified suitable continental proxy populations from 1000 genomes data and repeated the ADMIXTURE analysis in supervised mode. We labeled the three ancestral components as European (EUR), Amerindian (AMR), and African (AFR), based on similarity to the contemporary proxy populations giving a tri-partite compositional basis for each infant.

### 2.6 Non-Compositional Statistical Analysis

The study samples were chosen pragmatically based on availability of genome-wide genetic data and MPO measure(s) as described in 2.3. No prior sample size calculations were performed. Since the excreted MPO values ranged over multiple orders of magnitude, we log-transformed the raw MPO (‘the log rule’). Since neutrophils ^30, 31^ and MPO ^32, 33^ are also present in breastmilk, breast feeding status was an important covariate to consider. We stratified the Recodisa cases and controls because the cases were ascertained on a recent episode of diarrhea, reflecting enteric infection and immune response. Exploratory analysis of the MPO levels showed one very extreme outlier (1.7x value of the next most extreme) in the Recodisa study which was dropped. Except where noted, all statistical analysis was performed with R version 4.4.1 ^34^. The R car package (outlierTest) ^35^ was used to test for additional outliers in each study group at a Bonferroni corrected alpha level of 0.05. Outlier values in two other infants were winsorized to the next most extreme tail value. To estimate the variance explained by variation in genetic ancestry in the linear models, we compared R^2^ in the genetic ancestry null and genetic ancestry-tested models. In the MBC repeated measures model, we estimated the proportion of variance explained by the fixed effects by decomposing the variance components using the R2mlm R package ^36^.

To derive measures of socio-economic status (SES) for the infants, we retrieved variables on household income, ownership of durable household assets, and dwelling characteristics. Categorical variables with multiple responses/categories were recoded to binary (Yes/No) variables to include in the analysis. After dropping records with missing SES data we were left with 296 in Mal-ED Birth Cohort and 266 in Mal-ED Case Control. The Recodisa study did not capture these data. Principal component and factor analysis was performed separately for each study group using SPSS version 26.0 with eigenvalue greater than 1 and 25 maximum iterations of convergence. Direct oblique rotation (oblimin) rotation was chosen to exclude cases pairwise and suppress absolute correlation coefficients value < 0.3. PCA was performed separately for each cohort. The factor scores generated from first and second principal components were used as explanatory factors in the SES multiple regression models.

### 2.7 Compositional Analysis of Genetic Ancestry

Statistical analysis of compositional data is complicated by the simplex constraint of the data; for any single observation of a composition, the parts of the composition are constrained to be constant, so in this case for an individual genome (*i*):

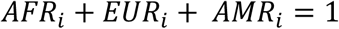

where *AFR_i_* (for example) was the fraction of African apportioned genetic ancestry in the *i*th infant. Here the compositional dimension is 3, usually symbolized with D in compositional statistics (equivalent to symbol K in genetic ancestry components). This constraint means that application of traditional statistics to such data, either descriptive to summarize the data, or to use as predictors in models, can give misleading and uninterpretable results ^19, 37, 38^. To measure the sample central tendency of the study groups we used the closed geometric mean:

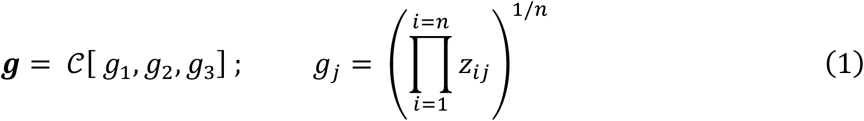

where *z_ij_* is the *jth* genetic ancestry for the *ith* infant and the closure operation is defined as:

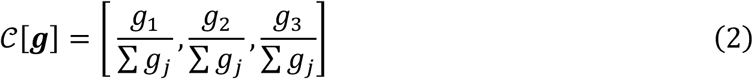

We compared sample variation of the genetic ancestry within study groups using the normalized variation matrix (T) which is a symmetric DxD matrix with zero diagonal elements, such that each element is given by:

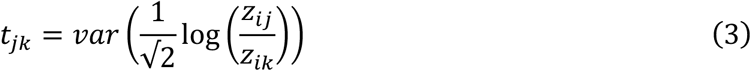

and the variance is the standard sample variance estimate. Each matrix entry directly measures the squared relative variability between pairs of genetic ancestries. Covariation of parts in a sample composition (correlation) also requires different approaches. Direct correlation analysis of the compositional parts leads to a negative bias and is only meaningful in a relative sense, such as in log ratio coordinates (below), but these coordinates have D-1 elements, thus excluding creation of an unequivocal D x D matrix ^39^. This has led to new work on correlation analysis of balances and sequential binary partitioning ^40^ but this is a complex subject and in lieu of this we used the elements of the variation matrix as measures of covariation ^19^. If *t_jk_* are near 0, the ratio is nearly constant and the two parts are almost proportional. Higher values of the log-ratio variance indicates high dispersion in the relative abundance and noisy proportionality between those two parts. The total variation of the composition sample is given by:

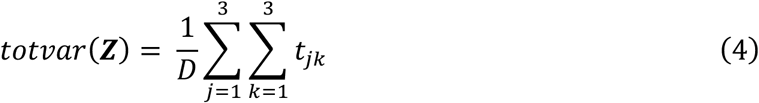

which serves as a scalar overall measure of dispersion of the data set in Aitchison geometry. In linear modeling using compositions as predictors, untransformed multivariate compositional parts are generally uninterpretable since it is impossible for a single part (eg *AFR_i_*) to vary independently with all other ancestral compositional parts held constant. From a linear analysis perspective, they are not a valid basis set for projection, since they are non-Euclidean. Hence to incorporate a compositional basis as predictors in a regression model, log ratios are most commonly used, reflecting the fact that composition parts vary relative to one another rather than absolutely. We used the additive log ratio (alr) oblique transformation ^41^:

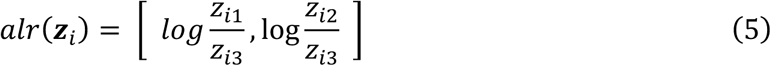

shown here with *z_i_*_3_ as the arbitrary reference; also the particular isometric log ratio (ilr) basis:

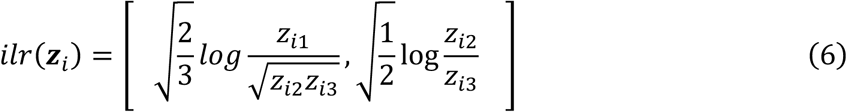

known as a pivot ilr orthogonal basis. Isometric log ratio transformation of the compositions creates an orthonormal basis of ratios that have the appropriate statistical properties ^42^. The pivot basis is a useful type of ilr because the first compositional part is isolated in the numerator of the first coordinate of *ilr*(***z****_i_*) allowing for interpretation of statistical inference results while the order of the other orthogonal components is irrelevant. In this situation, it measures the dominance of part *z_i_*_1_ with respect to the geometric average of the other parts. The pivot basis can be permuted into D (here D=3) separate isometric bases to test each of the three compositional parts as the first basis set predictor in three separate models ^43^.

Linear regression models for the MCC and the Recodisa groups were adjusted for infant gender, and age and breastfeeding status at fecal MPO sample time, hence the model with isometric log ratio transformed genetic ancestry composition was:

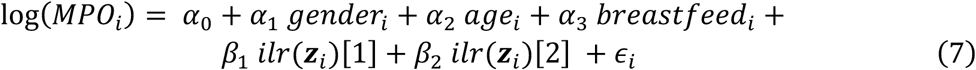

where ***z****_i_* = [ *AFR_i_*, *EUR_i_*, *AMR_i_*] or cyclically permuted tuples [ *EUR_i_*, *AMR_i_*, *AFR_i_*], [ *AMR_i_*, *AFR_i_*, *EUR_i_*]. An identical set of models was run replacing the ilr() transformation with the additive log ratio alr() for comparison. Inference was perfomed on the first term only, *ilr(**z**_i_)*[1] or *alr(**z**_i_)*[1]. The repeated measures models for MBC also included *age_i_*^2^ as a fixed effect and infant as a random intercept, and were run using the lme4 R package ^44^ with lmrTest ^45^. We tested inclusion of a random time-dependent slope for MBC under a genetic ancestry null model (no ilr or alr terms) but the random slope explained a trivial amount of additional variance, and therefore retained only random intercepts. Since the ilr and alr coordinates are interconvertible by linear transformation with a (D-1 x D-1) matrix, linear models in either coordinates give an identical result for an omnibus F-test of association of total genetic ancestry variation with the outcome, and partial R^2^ of trait variance explained by total genetic ancestry.

Interpretation of significant log ratio compositional association with an outcome is also more nuanced than often appreciated ^46^. A significant coefficient in a multiple linear regression model specifies that eg *β*_1_ reflects the estimated effect of ilr[1] (or alr[1]) on the expected log(MPO) *<u>assuming that all other model factors are constant, including ilr</u>*<u>[2] *(or alr*[2]).</u> Holding the second ilr term constant means that a change in z_1_ (eg AFR) has to be in the same proportion to z_2_ (eg EUR) *and* z_3_ (eg AMR) such that the ratio of z_2_/z_3_ is also constant. Thus the change in outcome comparing the first pivoted ilr variable for infant *h* (with genetic ancestry composition **z_h_**) vs infant *i* is:

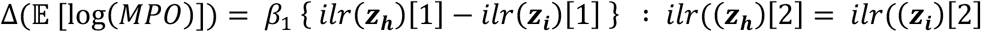

where *z*_ℎ_ is the 3-part composition vector for infant *h*. Rearranging:

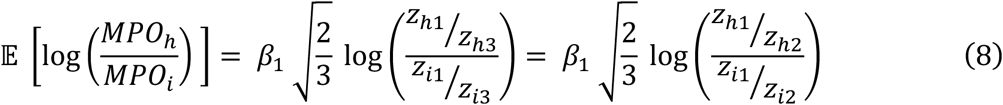

and this can be used to estimate the expected change in the log outcome given a contrast between two infant genetic ancestries that meet the z_2_/z_3_ constraint. Multiple linear models substituting an alr basis for ilr have an identical interpretation with the exception that normalizing constant √2/3 is absent. The ilr normalizing constants can be dropped and log ratios in base 2 or base 10 can also simplify interpretation of statistical linear model results. Since we have a log transformed outcome, only under a true lognormal distribution for MPO does:

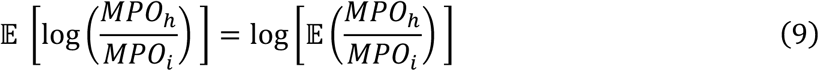

wherein the effect sizes can be transformed to an expected multiplier for the underlying MPO ratio, but in general, this does not hold, and if expression of the effect sizes as MPO ratios is needed, the delta method ^47^ to second order provides a more complex formula to correct the MPO level-dependent coefficient of variation. Since the goal of this study was primarily inferential, we used (9) to approximate the effect sizes as MPO ratios.

The ilr/alr transformation tests one particular class of compositional predictor hypotheses but a more natural question to ask is the effect of changing the ratio of two parts (eg AFR/EUR) while holding the third part (eg AMR) constant. A class of hypotheses of biological interest are those testing the effect of a change in sub-parts of ancestry while holding other parts constant, for example, the expected effect of a change +1% AFR, -1% EUR, or a change in %AFR/%EUR ratio while holding %AMR constant. We term this an isoancestry hypothesis (by analogy with isopleth diagrams). This type of hypothesis is not readily amenable to log ratio models and is emblematic of a more general criticism of log ratio bases; while rigorous linear algebra emphasizes orthogonality and hence minimizes collinearity, the results are difficult to interpret^38^. To preserve the rigor of the log ratio models but enable this class of hypothesis to be tested, we developed a new post-estimation analysis method using average marginal effects. Generalizing (7) using additive log ratios (alr), with W_r_ as the *r*th covariate random variable and Z_k_as the random variable for part *k* of the composition:

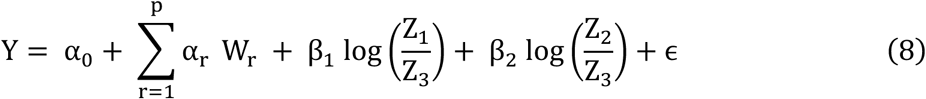

Imposing the constraint that eg part 2 is held constant, Z_2_ = C, and after differentiating and simplifying:

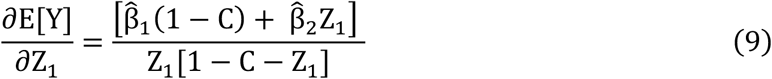

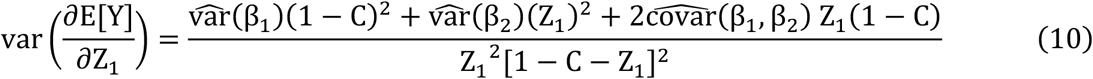

and since:

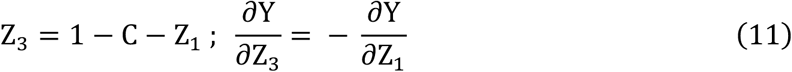

the corresponding Z_3_ estimates are easily derived. Computing (9) and (10) for each individual in a data set and then averaging gives an estimate of the expected marginal change in outcome Y, holding Z_2_constant, as Z_1_increases by some unit with an obligate decrease in Z_3_ by the same amount. This is a linear shift isoancestry test that provides an estimate of the effect and an hypothesis test H_0_ : 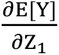 = 0. Similarly, defining the log ratio of the two varying parts:

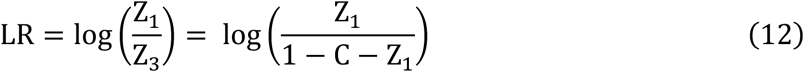

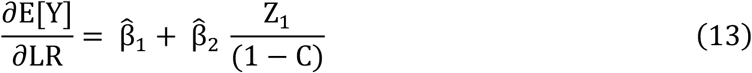

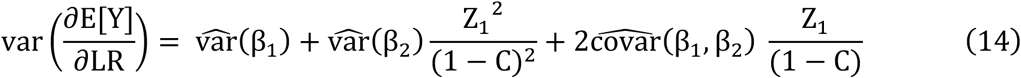

which provides a corresponding log ratio shift isoancestry estimate and test. These formulas are easily generalizable to D>3 part genetic ancestry compositions where D-2 parts are held constant and are combined into the overall constant C.

## Results

### 3.1 Study Populations

We assembled a complete data set of 945 infants with genetic ancestry, covariate, and fecal MPO biomarker measures, drawn from three pediatric studies (four study groups) recruited from within four states in Northeastern Brazil (Table 1 and Figure 1A). These included 243 from the Brazil site of the Mal-ED Birth Cohort (MBC); 289 from the corresponding Mal-ED Case Control Cohort (MCC); 415 from the Recodisa Case-Control Study stratified into separate case and control groups. Notably, a variable but quite large proportion of mothers were still breastfeeding to some degree at infant age 1-2 years. The highest median fecal MPO levels were in the Recodisa Cases and MCC groups, and infants in the latter study also had the poorest anthropometry HAZ/WAZ scores. The stools in the 4-state REC study were collected at a mean of about 22 months of age, while the MBC and MCC studies (conducted in Fortaleza) were collected closer to one year of age. The stool MPO values varied across two orders of magnitude (Table 1), with the highest median and greatest variation in the REC Cases, followed by the MCC study. The log(MPO) distributions were all negatively skewed (Figure 1B), and were almost bimodal in the MBC and REC-Co groups with an emerging peak at lower log(MPO) (between 0-5 log ng/nL)

**Figure 1.**
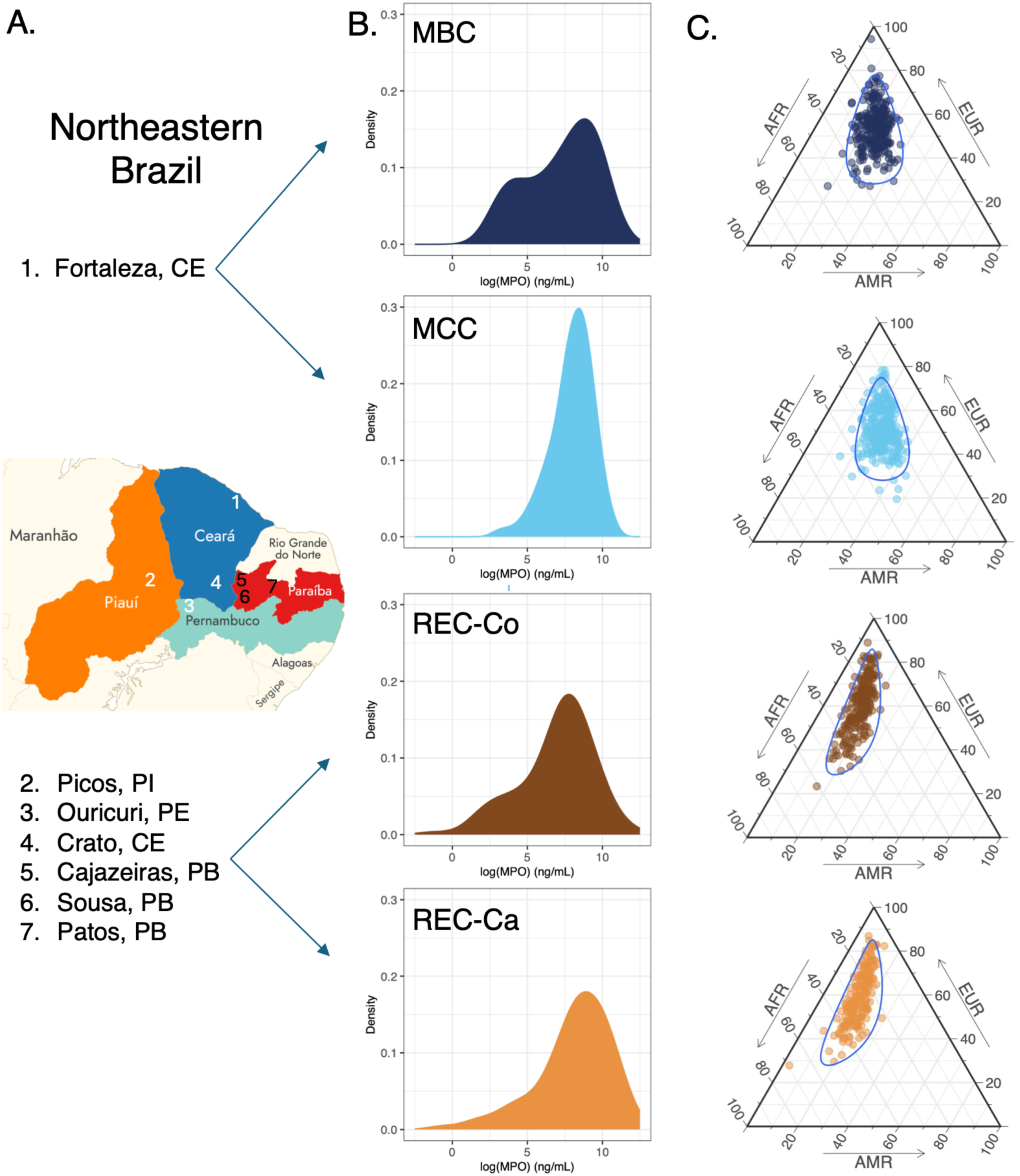
Geographical and study sources of infant fecal samples, their distribution of myeloperoxidase (MPO), and continental genetic ancestry. A. Cities (states) in Northeastern Brazil where infants were enrolled into the studies. B. For each study group, the distribution of log(MPO) (ng/mL). MBC: Mal-ED Birth Cohort; MCC: Mal-ED Case Control; REC-Co: Recodisa Study Controls; REC-Ca: Recodisa Study Cases. C. For each study group, a ternary plot of tripartite continental genetic ancestry (%AFR, %EUR, %AMR). Each colored dot is an infant in that study. Top vertex corresponds to 100% EUR. Left vertex 100% AFR. Right vertex 100% AMR.

**Table 1.**
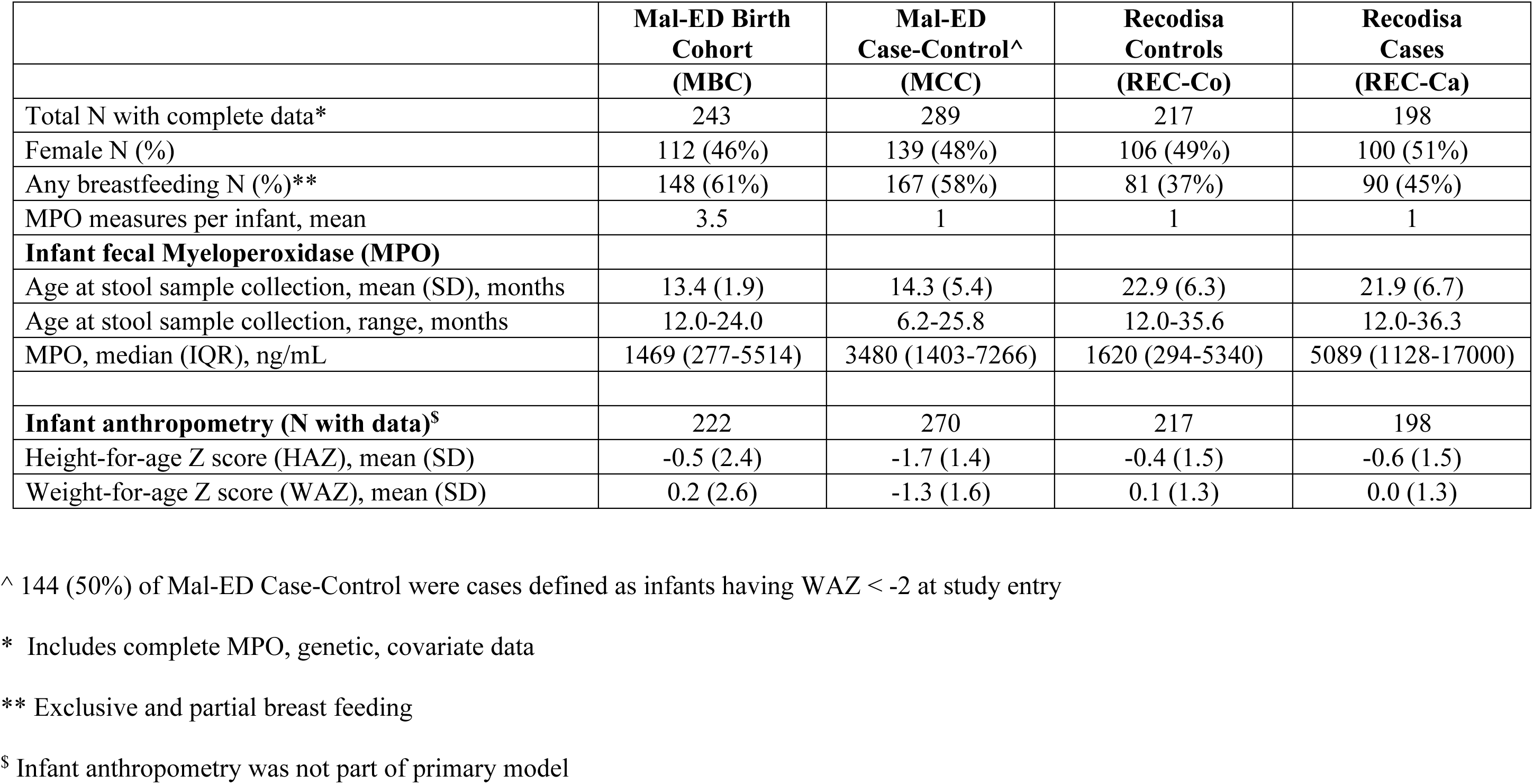
Clinical characteristics of the infants of the four Northeastern Brazil study groups with fecal MPO measures.

### 3.2 Genetic Ancestry Compositional Analysis

The study groups included admixed Brazilian infants with estimated major continental tripartite genetic African, European, and Amerindian ancestry. The composition varied amongst the study groups (Table 2 and Figure 1C). All 4 of the included study groups had a mean (centre) African genetic ancestry of about 22-25% but samples drawn from the smaller and more rural sites in Recodisa carried greater European and lower Amerindian (means about 60% and 14% respectively) versus the urban Fortaleza study infants with lower European and higher Amerindian (means 24% and 22%). The study group ternary plots in Figure 1C visually show the distinction between the Fortaleza and more rural sites in the distribution of continental genetic ancestry. Of the three pairs of ancestries, African and European were the least ‘correlated’ having the highest normalized variance of the log ratio in all study groups. Conversely, African and Amerindian were the most ‘correlated’ (value closest to zero). The variation of ancestry was greater in the REC groups compared to the Fortaleza studies (MBC and MCC), as measured by the range in fractional ancestry and the total normalized variation (Table 1). Contrary to recent previous assertions that sampled Brazilian Amerindian ancestry in genome-wide studies has been limited to less than 10% ^48^, the range of Amerindian ancestry extended to above 30% and 25% respectively, in the Fortaleza and Recodisa studies.

**Table 2.** Summary statistics for infant continental genetic ancestry composition, by study group.

|  | **Mal-ED Birth Cohort (MBC)** | **Mal-ED Case-Control (MCC)** | **Recodisa Controls (REC-Co)** | **Recodisa Cases (REC-Ca)** |
| --- | --- | --- | --- | --- |
| Study Group Sample (N) | 243 | 289 | 217 | 198 |
| **Genetic Ancestry Fraction** |  |  |  |  |
| f(AFR), range | 0.04 - 0.55 | 0.09 - 0.46 | 0.07 – 0.61 | 0.05 – 0.69 |
| f(EUR), range | 0.27 - 0.94 | 0.20 – 0.78 | 0.23 – 0.89 | 0.28 – 0.87 |
| f(AMR), range | 0.02 - 0.40 | 0.11 – 0.47 | 0.03 – 0.26 | 0.03 – 0.28 |
| **Genetic Ancestry Centre\*** |  |  |  |  |
| f(AFR) [95% CI] | 0.22 [0.08, 0.36] | 0.23 [0.11, 0.35] | 0.24 [0.05, 0.43] | 0.25 [0.05, 0.45] |
| f(EUR) [95% CI] | 0.54 [0.35, 0.74] | 0.52 [0.33, 0.71] | 0.61 [0.37, 0.86] | 0.60 [0.36, 0.85] |
| f(AMR) [95% CI] | 0.24 [0.12, 0.35] | 0.25 [0.12, 0.38] | 0.14 [0.06, 0.23] | 0.14 [0.06, 0.23] |
| f(Total)** | 1.00 | 1.00 | 1.00 | 1.00 |
| **Normalized Variance of Log Ratio** |  |  |  |  |
| log(AFR/EUR) | 0.121 | 0.103 | 0.194 | 0.201 |
| log(AFR/AMR) | 0.064 | 0.055 | 0.061 | 0.080 |
| log(EUR/AMR) | 0.106 | 0.098 | 0.080 | 0.114 |
| **Total Normalized Variation** | 0.194 | 0.170 | 0.255 | 0.263 |

\* Centre is the closed geometric mean within a study group and is a measure of central tendency (mean). For historical reasons this is typically written with British English spelling.
\*\* Differences from column sums are due to rounding

### 3.3 Relative African and European Ancestries are Associated with Quantitative Level of Excreted Fecal MPO

We tested the association of estimated genetic ancestry of admixed Brazilian infants with fecal log(MPO) levels to establish whether there could be a genetic influence on the biomarker. We sequentially and separately tested the association of the dyadic basis coordinates in ilr() and alr() as variables appropriate for the compositional nature of genetic ancestry proportions adjusting for gender, age and breastfeeding status at study sample timepoint (Table 3).

**Table 3.**
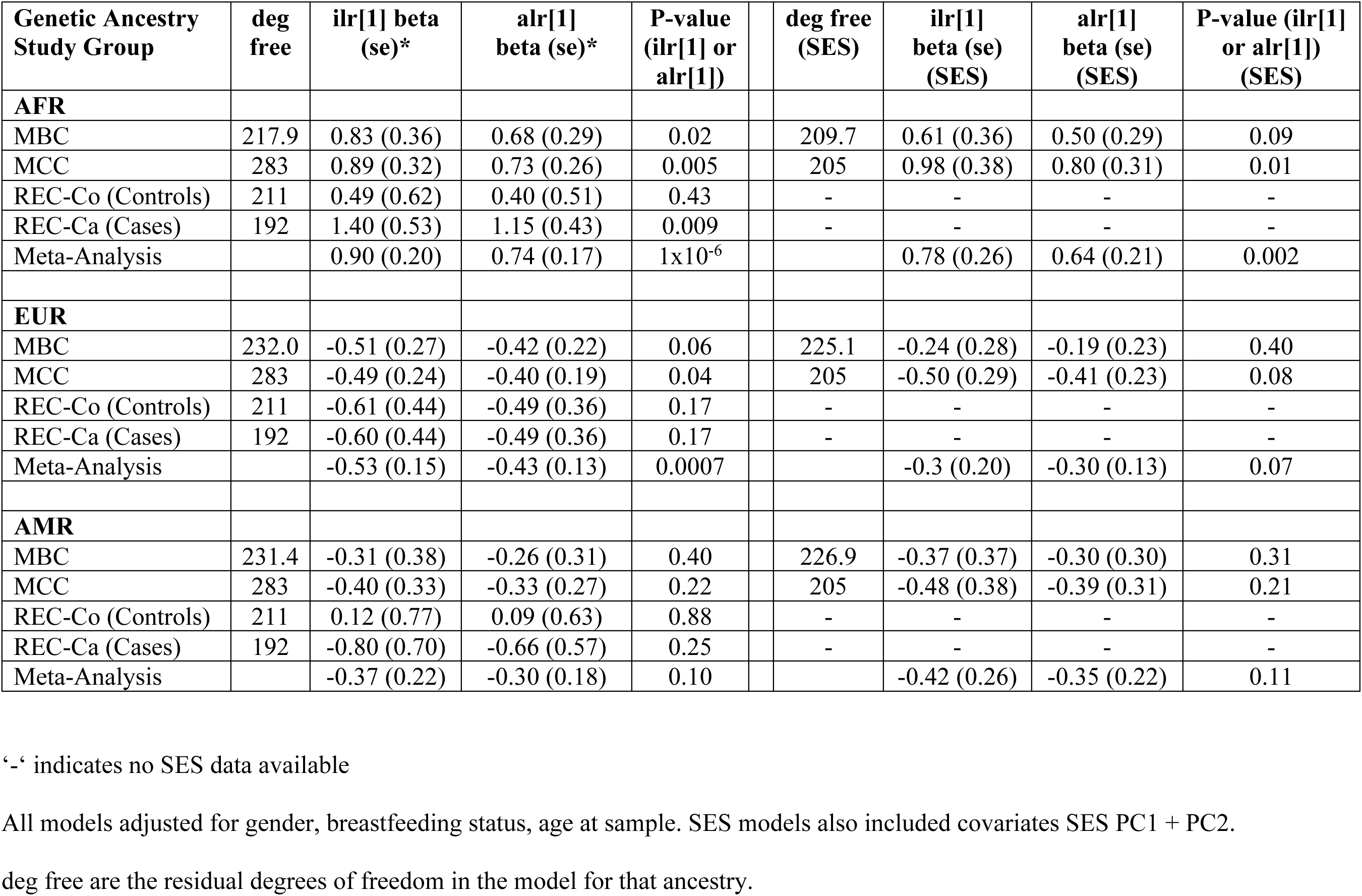
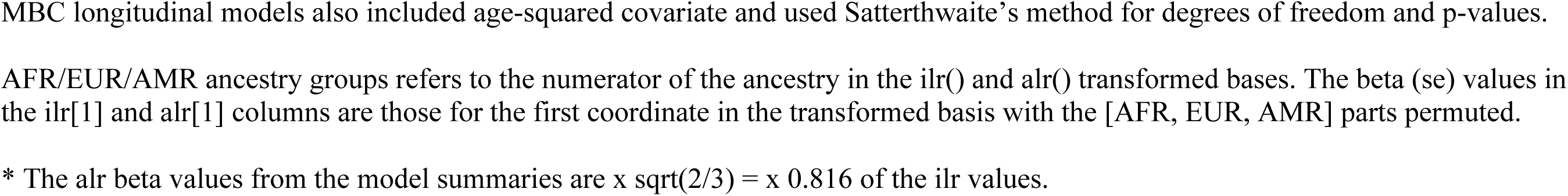
Tests of association of continental genetic ancestry with fecal log(MPO) in 4 Brazil study groups, models without and with SES components.

| **Genetic Ancestry Study Group** | **deg free** | **ilr[1] beta (se)\*** | **alr[1] beta (se)\*** | **P-value (ilr[1] or alr[1])** | **deg free (SES)** | **ilr[1] beta (se) (SES)** | **alr[1] beta (se) (SES)** | **P-value (ilr[1] or alr[1]) (SES)** |
| --- | --- | --- | --- | --- | --- | --- | --- | --- |
| **AFR** |  |  |  |  |  |  |  |  |
| MBC | 217.9 | 0.83 (0.36) | 0.68 (0.29) | 0.02 | 209.7 | 0.61 (0.36) | 0.50 (0.29) | 0.09 |
| MCC | 283 | 0.89 (0.32) | 0.73 (0.26) | 0.005 | 205 | 0.98 (0.38) | 0.80 (0.31) | 0.01 |
| REC-Co (Controls) | 211 | 0.49 (0.62) | 0.40 (0.51) | 0.43 | - | - | - | - |
| REC-Ca (Cases) | 192 | 1.40 (0.53) | 1.15 (0.43) | 0.009 | - | - | - | - |
| Meta-Analysis |  | 0.90 (0.20) | 0.74 (0.17) | 1x10<sup>-6</sup> |  | 0.78 (0.26) | 0.64 (0.21) | 0.002 |
| **EUR** |  |  |  |  |  |  |  |  |
| MBC | 232.0 | -0.51 (0.27) | -0.42 (0.22) | 0.06 | 225.1 | -0.24 (0.28) | -0.19 (0.23) | 0.40 |
| MCC | 283 | -0.49 (0.24) | -0.40 (0.19) | 0.04 | 205 | -0.50 (0.29) | -0.41 (0.23) | 0.08 |
| REC-Co (Controls) | 211 | -0.61 (0.44) | -0.49 (0.36) | 0.17 | - | - | - | - |
| REC-Ca (Cases) | 192 | -0.60 (0.44) | -0.49 (0.36) | 0.17 | - | - | - | - |
| Meta-Analysis |  | -0.53 (0.15) | -0.43 (0.13) | 0.0007 |  | -0.3 (0.20) | -0.30 (0.13) | 0.07 |
| **AMR** |  |  |  |  |  |  |  |  |
| MBC | 231.4 | -0.31 (0.38) | -0.26 (0.31) | 0.40 | 226.9 | -0.37 (0.37) | -0.30 (0.30) | 0.31 |
| MCC | 283 | -0.40 (0.33) | -0.33 (0.27) | 0.22 | 205 | -0.48 (0.38) | -0.39 (0.31) | 0.21 |
| REC-Co (Controls) | 211 | 0.12 (0.77) | 0.09 (0.63) | 0.88 | - | - | - | - |
| REC-Ca (Cases) | 192 | -0.80 (0.70) | -0.66 (0.57) | 0.25 | - | - | - | - |
| Meta-Analysis |  | -0.37 (0.22) | -0.30 (0.18) | 0.10 |  | -0.42 (0.26) | -0.35 (0.22) | 0.11 |

‘-’ indicates no SES data available
All models adjusted for gender, breastfeeding status, age at sample. SES models also included covariates SES PC1 + PC2.
deg free are the residual degrees of freedom in the model for that ancestry.
MBC longitudinal models also included age-squared covariate and used Satterthwaite's method for degrees of freedom and p-values.
AFR/EUR/AMR ancestry groups refers to the numerator of the ancestry in the `ilr()` and `alr()` transformed bases. The beta (se) values in the `ilr[1]` and `alr[1]` columns are those for the first coordinate in the transformed basis with the [AFR, EUR, AMR] parts permuted.
\* The alr beta values from the model summaries are $\times \sqrt{2/3} = \times 0.816$ of the ilr values.

We confirmed that the association beta/se results for ilr() and alr() only differed by the normalizing ilr constant in the first coordinate (sqrt(2/3)) with identical P-value and outcome variance explained by the total genetic ancestry variance. Using the pivoted ilr() coordinates tested in 3 separate models for each study group and then after meta-analyzing, we found that an increase in the proportion of AFR ancestry relative to a fixed EUR/AMR ancestry ratio was associated with an increase in MPO levels (alr[1] beta= +0.74, P=1x10^-6^). This was present with a smaller secondary association of decrease of MPO levels with an increasing relative EUR ancestry while holding the AMR/AFR ratio constant (alr[1] beta=-0.43, P=7x10^-4^). There was no evidence for association of change in relative AMR (holding EUR/AFR constant) with MPO (P=0.10). Utilizing equation (8) (Methods), for a doubling of AFR/EUR ancestry ratio, the meta-analysis point estimate for mean change in log(MPO) was 0.74 x log(2) = +0.51 for an approximate expected 1.7x increase in MPO. Similarly, for a relative change in EUR ancestry proportion to AMR holding AFR/AMR constant (or vice versa) the point estimate was -0.3 in the mean change in log(MPO) for an expected 0.7x decrease in MPO.

Since genetic ancestry is heavily confounded with race and hence SES, we developed principal component scores to quantitatively measure the SES environment of the infants as additional model adjustments, including variables based on household income, ownership of durable household assets, and dwelling characteristics. We repeated the analyses for the two studies with socio-economic family data and included two SES principal component scores as adjustments (Table 3). We found similar results to non-SES adjusted data, albeit with a much smaller sample size, but with attenuation of the genetic ancestry effects. The association of relative AFR ancestry with log(MPO) was still significant (P=0.002), while the negative association of relative EUR ancestry was no longer significant but only marginally missed the alpha threshold (P=0.07).

### 3.4 Infant Fecal MPO Variance Explained by Differences in Genetic Ancestry

Using the ilr and alr linear models, we estimated the variance explained by total genetic ancestry by measuring the incremental increase in variance explained in models with and without the genetic ancestry factors. The explained variance was found to be quite modest, ranging from 1.2% in Recodisa controls to 4.3% in Recodisa cases (Table 4) but the greater variance in the Recodisa cases and attenuation in controls suggested a possible operative selection process. Since the Recodisa cases were ascertained on occurrence of recent diarrheal episodes within 24 hours of enrollment, if continental differential genetic selection has occurred, it has operated to modify the innate neutrophil *response* to enteric infection rather than the prevalent background lower-level MPO excretion during non-infected periods. The MCC study group of infants showed the second highest variance explained (3.1%) consistent with the enteric pathogen load found in their surveillance stool samples using TaqMan PCR methods^21^. The pathogen frequency results suggested a fraction of about 20% of these infants could have been study recruited during an enteric infection episode.

**Table 4.** Fecal log(MPO) variance explained by genetic factors for different models, without and with SES adjustment.

| **Study Group** | **Ilr** | **alr** | **Untrans-**<br>**formed** |  | **Ilr**<br>**(SES)** | **alr**<br>**(SES)** | **Untrans-**<br>**formed (SES)** |
| --- | --- | --- | --- | --- | --- | --- | --- |
| MBC | 1.4 % | 1.4 % | 1.6 % |  | 0.5 % | 0.5 % | 0.7 % |
| MCC | 3.1 % | 3.1 % | 2.6 % |  | 3.3 % | 3.3 % | 2.8 % |
| REC-Co (Controls) | 1.2 % | 1.2 % | 1.2 % |  | - | - | - |
| REC-Ca (Cases) | 4.3 % | 4.3 % | 3.7 % |  | - | - | - |

‘-‘ indicates no SES data available
Outcome variance explained by total genetic ancestry variation in multiple linear models with: 1) joint ilr[1] + ilr[2] coordinates and similarly 2) joint alr[1] and alr[2]); 3) Untransformed: with $ANC_1 + ANC_2$ untransformed for comparison.

For comparison we ran the same models but substituted two untransformed independent parts of the three-part ancestry composition rather than log ratios (any two of the three give the same results). The fraction of variance explained by genetic ancestry factors was comparable or generally slightly reduced, although the individual ancestry factor associations were highly collinear with large variance inflation factors, and were uninterpretable per se. The differences in total variance explained is due to the subtleties of the correlations between the log ratios or the untransformed genetic proportions vis-a-visa the other included model covariates. In the two study groups that had captured SES data, the variance explained in the SES PC adjusted models was either reduced by about 50% (MBC) to 0.5% variance explained; or unchanged (MCC) with 3.3% explained, suggesting that some components of the genetic ancestry association may be independent of SES and hence race.

### 3.5 Translation of Log Ratios into Tests of Isoancestry Hypotheses

The log ratio model results described in section 3.3 are rigorous and have a coherent interpretation rooted in linear algebra, but that interpretation remains convoluted and non-intuitive for genetic admixing processes. We developed a method to test hypotheses with an isoancestry constraint using post-estimation average marginal effects. This approach enables two new classes of hypotheses to be tested. The isoancestry linear shift tests the mean change in outcome given a predictor contrast of {Z_1_ + X %, Z_3_ - X%, Z_2_ : Constant} while the isoancestry log ratio tests {log(Z_1_ / Z_3_) = +X, Z_2_ : Constant}. Figure 2 gives a geometric comparison and explanation of these hypotheses. We ran these tests for all study groups and all permuted genetic ancestries, without and with SES adjustments. Figure 3 shows the forest diagrams summarizing the results of these tests compared with alr() results from Table 3 (rescaled to doubling of predictor ratio), while Table 5 shows the numerical details of the results.

**Figure 2.**
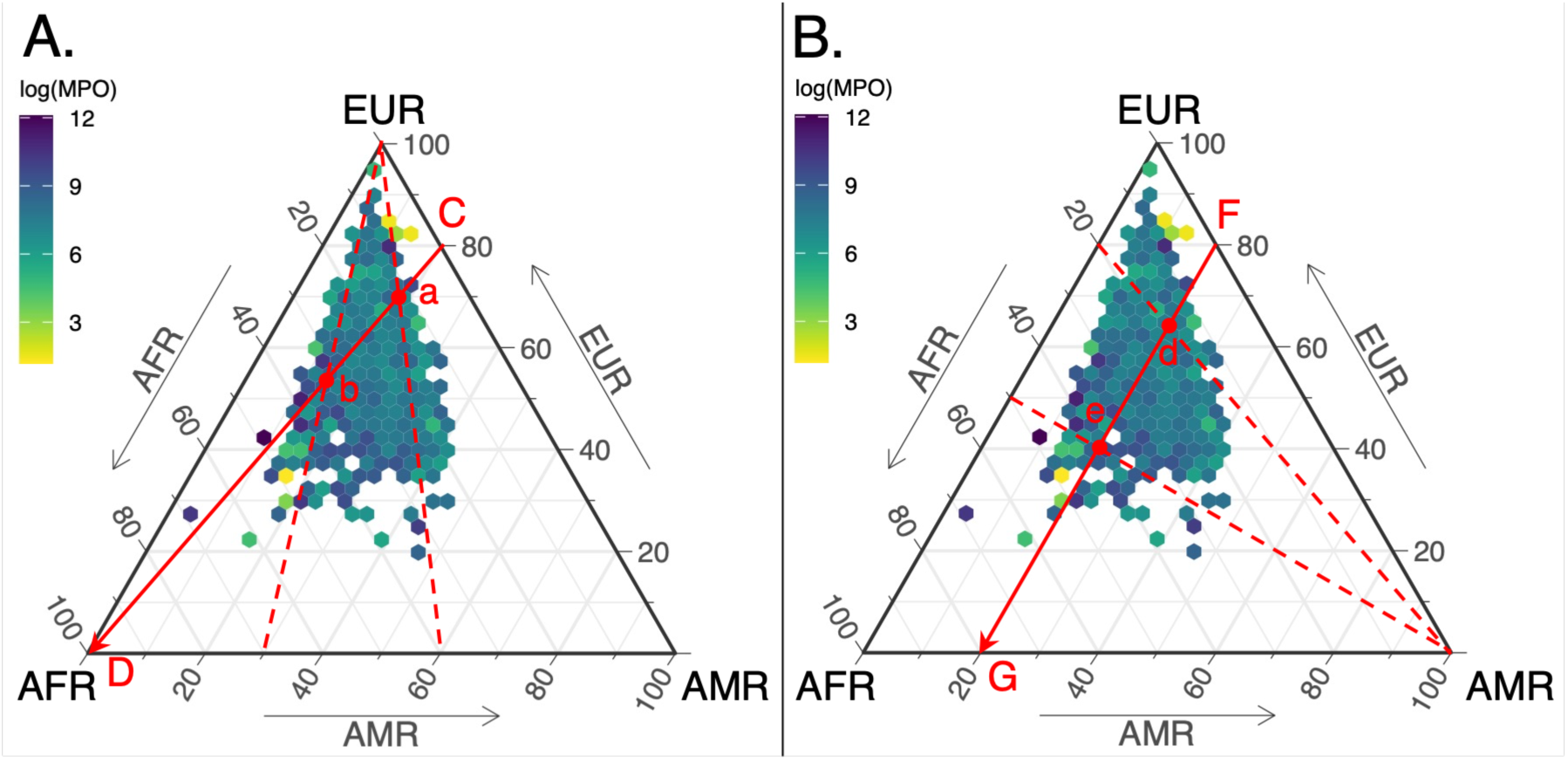
Ternary hexagonal density diagrams illustrating the hypotheses being tested in traditional log ratio and isoancestry models. Small hexagons are colored by the mean log(MPO) for infants that fall within that local ancestry composition range. Genetic ancestry and MPO outcome data for all four infant study groups are combined in each plot. Vertices labeled AFR, EUR, and AMR are the points of 100% ancestry for each of these continental groups. Edge arrows show the direction of increasing ancestry for that continent. Although noisy, a general trend of increasing log(MPO) towards the lower left AFR vertex is visible. The hexagon distribution is identical in A. and B. A. Geometric interpretation of log ratio tests (ilr and alr are equivalent here). Along the example red cevian line segment CD the ratio of EUR / AMR ancestry remains constant at 80/20=4/1. The relative fraction of AFR ancestry increases from C (0%) to D (100%). Points a and b (in red) are example compositional ratios (EUR/AMR, AFR /AMR) a.(80/20, 40/60) giving AFR=12% and b. (80/20, 70/30), AFR=32%, respectively. The first ilr basis set genetic log ratio variable ilr[1] = log(AFR/sqrt(EUR*AMR)) estimates the mean change in log(MPO) in all cevian segments ending at D across the domain of all possible fixed EUR/AMR ratios seen in the data. Since ilr[2] is held constant in the regression interpretation of ilr[1], log(EUR/AMR) is constant and alr[1] = log(AFR/AMR) has an identical interpretation. B. Geometric interpretation of the isoancestry models. Along the example red isoancestry line segment FG, AMR genetic ancestry is fixed at 20%. The fraction of AFR ancestry increases from 0% at F to the maximum possible 80% at G. Concomitantly, the ratio of AFR/EUR increases from 0/80 at F to 80/0 at G. Points d and e are examples ratios on FG, respectively with (AFR/EUR) = 20/80 (1/4) and 50/50 (1/1). Isoancestry models holding AMR constant test the mean change in log(MPO) along all fixed AMR isoancestry line segments that overlap the domain of the fractional AMR ancestries seen in the data. Similarly for isoancestries for the other two continents.

**Figure 3.**
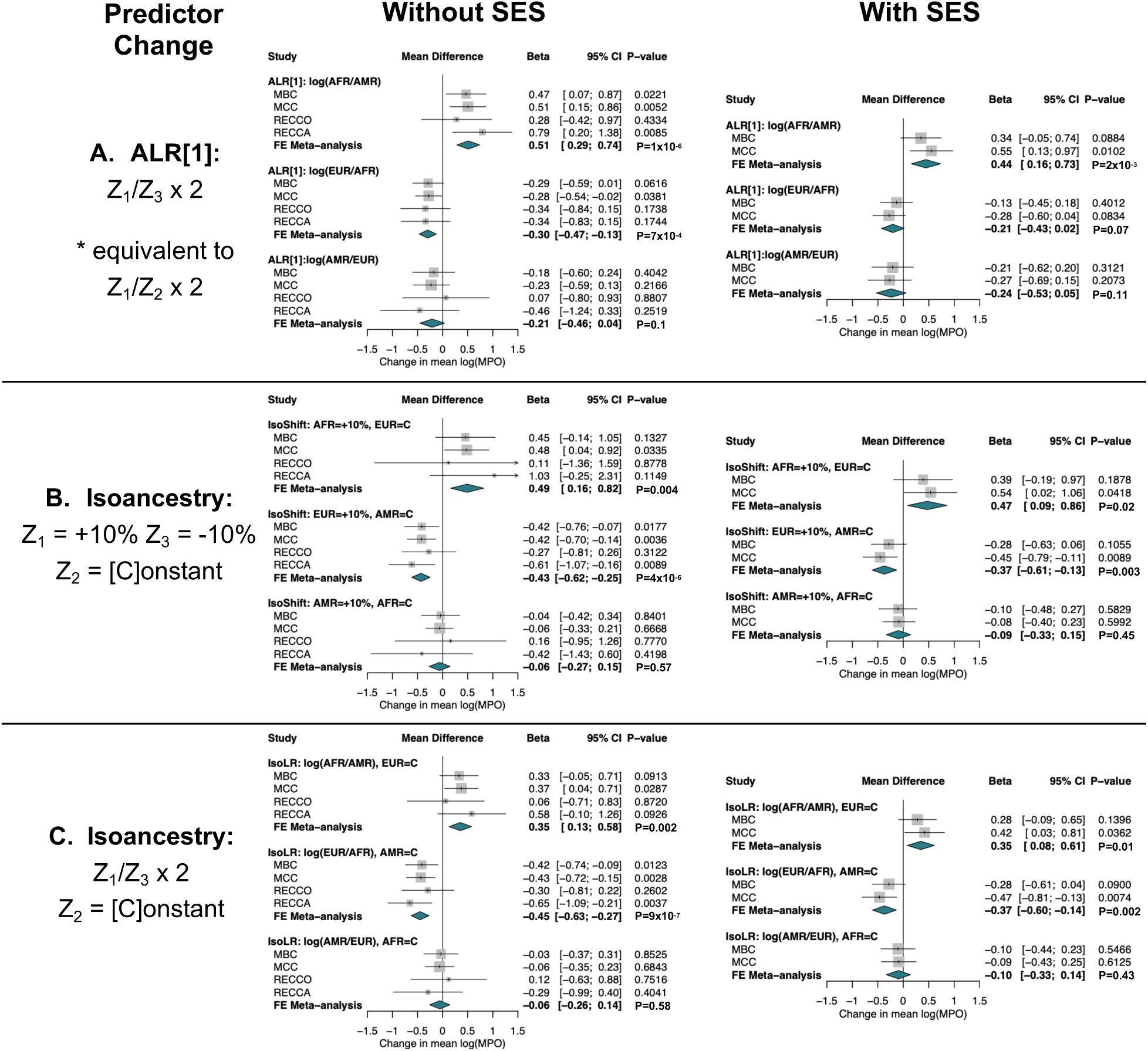
Forest plots summarizing the individual study association results of ancestry with log(MPO) for three classes of test, without and with socio-economic status (SES) adjustments. Left column of forest plots (Without SES) did not include SES PC covariates. Right column (With SES) included SES principal component 1 and 2. Left Predictor Change column specifies the change in the predictor for which beta (the estimated model coefficient for that predictor) measures the change in outcome, mean log(MPO). A. (top row). ALR[1] is the additive log ratio basis for the first variable in the ALR basis. ALR[1]: predictor change is Z_1_/Z_3_ ratio doubling with log(Z_1_/Z_3_) held constant. These beta and se values are multiplied (rescaled) by log(2) = 0.693 compared to the values in Table 3. B. (middle row) IsoShift is isoancestry with linear shift variable. Predictor change is +10% Z_1_ -10% Z_3_, with Z_2_ held constant. C. (bottom row) IsoLR is isoancestry with log ratio variable. Predictor change is for Z_1_/Z_3_ ratio doubling with Z_2_ held constant.

**Table 5.**
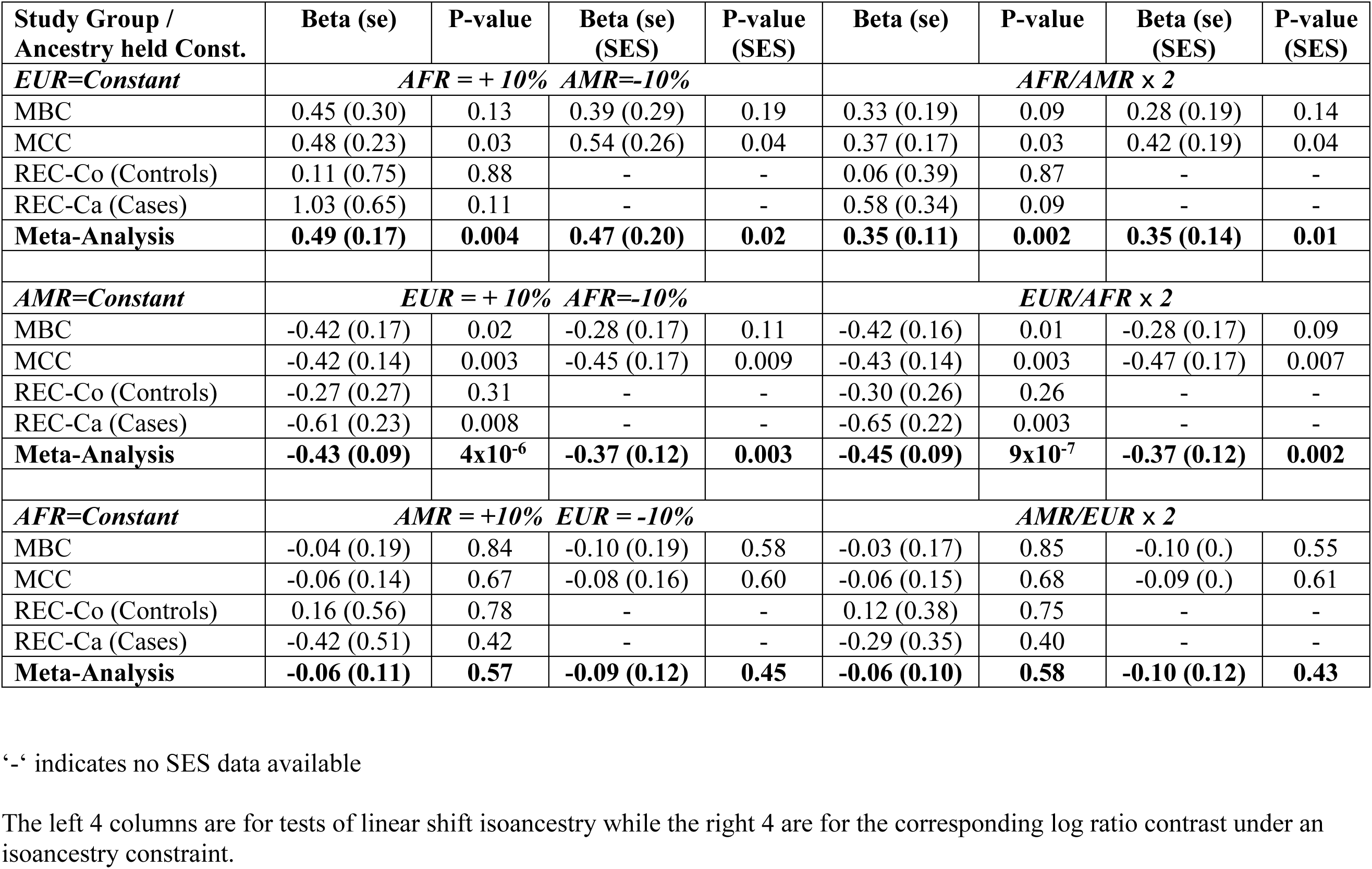
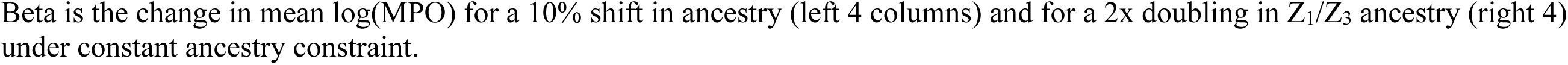
Tests of association of continental genetic ancestry with infant fecal log(MPO) using tests under isoancestry hypotheses.

| Study Group / Ancestry held Const. | Beta (se) | P-value | Beta (se) (SES) | P-value (SES) | Beta (se) | P-value | Beta (se) (SES) | P-value (SES) |
| --- | --- | --- | --- | --- | --- | --- | --- | --- |
| ***EUR=Constant*** | ***AFR = + 10% AMR=-10%*** |  |  |  | ***AFR/AMR × 2*** |  |  |  |
| MBC | 0.45 (0.30) | 0.13 | 0.39 (0.29) | 0.19 | 0.33 (0.19) | 0.09 | 0.28 (0.19) | 0.14 |
| MCC | 0.48 (0.23) | 0.03 | 0.54 (0.26) | 0.04 | 0.37 (0.17) | 0.03 | 0.42 (0.19) | 0.04 |
| REC-Co (Controls) | 0.11 (0.75) | 0.88 | - | - | 0.06 (0.39) | 0.87 | - | - |
| REC-Ca (Cases) | 1.03 (0.65) | 0.11 | - | - | 0.58 (0.34) | 0.09 | - | - |
| **Meta-Analysis** | **0.49 (0.17)** | **0.004** | **0.47 (0.20)** | **0.02** | **0.35 (0.11)** | **0.002** | **0.35 (0.14)** | **0.01** |
| ***AMR=Constant*** | ***EUR = + 10% AFR=-10%*** |  |  |  | ***EUR/AFR × 2*** |  |  |  |
| MBC | -0.42 (0.17) | 0.02 | -0.28 (0.17) | 0.11 | -0.42 (0.16) | 0.01 | -0.28 (0.17) | 0.09 |
| MCC | -0.42 (0.14) | 0.003 | -0.45 (0.17) | 0.009 | -0.43 (0.14) | 0.003 | -0.47 (0.17) | 0.007 |
| REC-Co (Controls) | -0.27 (0.27) | 0.31 | - | - | -0.30 (0.26) | 0.26 | - | - |
| REC-Ca (Cases) | -0.61 (0.23) | 0.008 | - | - | -0.65 (0.22) | 0.003 | - | - |
| **Meta-Analysis** | **-0.43 (0.09)** | **4x10<sup>-6</sup>** | **-0.37 (0.12)** | **0.003** | **-0.45 (0.09)** | **9x10<sup>-7</sup>** | **-0.37 (0.12)** | **0.002** |
| ***AFR=Constant*** | ***AMR = +10% EUR = -10%*** |  |  |  | ***AMR/EUR × 2*** |  |  |  |
| MBC | -0.04 (0.19) | 0.84 | -0.10 (0.19) | 0.58 | -0.03 (0.17) | 0.85 | -0.10 (0.) | 0.55 |
| MCC | -0.06 (0.14) | 0.67 | -0.08 (0.16) | 0.60 | -0.06 (0.15) | 0.68 | -0.09 (0.) | 0.61 |
| REC-Co (Controls) | 0.16 (0.56) | 0.78 | - | - | 0.12 (0.38) | 0.75 | - | - |
| REC-Ca (Cases) | -0.42 (0.51) | 0.42 | - | - | -0.29 (0.35) | 0.40 | - | - |
| **Meta-Analysis** | **-0.06 (0.11)** | **0.57** | **-0.09 (0.12)** | **0.45** | **-0.06 (0.10)** | **0.58** | **-0.10 (0.12)** | **0.43** |

‘-’ indicates no SES data available
The left 4 columns are for tests of linear shift isoancestry while the right 4 are for the corresponding log ratio contrast under an isoancestry constraint.
Beta is the change in mean $\log(\text{MPO})$ for a 10% shift in ancestry (left 4 columns) and for a 2x doubling in $Z_1/Z_3$ ancestry (right 4) under constant ancestry constraint.

Overall, substituting EUR for AFR ancestry at constant AMR was associated with increased fecal MPO (meta-analysis beta=0.43 for 10% shift of EUR to AFR, P=4x10^-6^). Similarly, a 10% shift of AMR to AFR ancestry at constant EUR was associated with a smaller effect (beta=0.49, P=0.004). There was no association for the corresponding shift from EUR to AMR ancestry at constant AFR (P=0.57). The corresponding tests under the isoancestry log shift test were both more significant (P=9x10^-7^ and P=0.002 respectively) suggesting that the log ratio contrast under constant AMR and constant EUR constraints had the greatest statistical power of all of the tests applied. This is likely highly dependent on the distribution of the ancestry compositions and outcome data sampled in the project, and may not be true for other outcomes or ancestry compositions. The trends in the results remained for models with additional SES covariate adjustment although with some attenuation of effect size and reduced significance due to reduced sample size.

## Conclusions

Fecal MPO is of interest as a biomarker of enteric immune response, inflammation, and pathogen clearance. We applied compositional analysis to the admixture proportions of major continental ancestry in Brazilian infants, including African, European and Amerindian, to test whether fecal MPO as a biomarker systematically varies with genetic ancestry, and hence gather possible evidence for a genetic explanatory component. Using log ratio ilr() and alr() bases we showed that increase in the relative proportion of AFR ancestry with fixed EUR/AMR ratio is associated with increased excreted MPO, with a secondary smaller negative association of EUR ancestry relative to fixed AMR/AFR ratio. To enable testing under a fixed isoancestry constraint we developed a new post-estimation average marginal effect method that uses estimates from fitting the orthogonal genetic ancestry compositional log ratio basis, enabling inferences under a constrained ancestry part with a linear shift or log ratio contrast. We found that a shift of EUR to AFR ancestry at constant fractional AMR was associated with increased fecal MPO, with a smaller secondary increase when for substituting AMR ancestry for AFR at constant EUR. Overall, this suggests that African genomes carry an additional aggregation of alleles that enable a greater neutrophilic response to enteric disease as measured by excreted MPO, perhaps under some specific selective environment. While the variance explained by ancestry differences in the MPO biomarker outcome was quite modest (1-4%), the study groups likely carrying the highest enteric pathogen burden at fecal sampling had the highest variance explained. Also, the variance explained in this context measures genetic ancestral differences in heritability, not the absolute heritability. Inclusion of covariates measuring SES into the models did not fully attenuate the effects providing some evidence that these associations reflect underlying genetics rather than SES (and indirectly, race), but caution is necessary in interpreting these results. Finally, although we have discussed these fractional ancestry analytical considerations in a global ancestry analysis, they may be pertinent in local ancestry analyses also, dependent on the how the ancestry is modeled.

In many previous papers that tested the association of genetic ancestry composition with an outcome biomarker or disease, the fractional composition has been analyzed as untransformed fractional variables because geneticists may not be aware of the problems of interpreting effects from raw compositional parts^49–54^. Although the D total parts of a genetic ancestry (often labeled K in genetic ancestry analysis) are overdetermined, merely entering D-1 parts that vary independently does not address the underlying issues of the inherent collinearity of the ancestry fractions, the large variance inflation factors, and the problem in interpreting the coefficients in a standard multiple linear model. Restricting models to a single fractional predictor can also lead to false conclusions if the composition is multi-part and the apparent association is due to a component summarized in the denominator. Researchers dealing with microbiome OTUs, cell type fractions in transcriptomics, or sample measured cell types in immunology are increasingly aware of these analytical issues ^38, 55, 56^. Another approach to handling genetic ancestry is to classify individuals into a small number of categorical genetic ancestry groups that are assumed homogeneous, using a clustering approach based on the primary continental principal components analysis^57–59^. While this removes the compositional complexities, the disadvantage is that it ignores the empirical continuous admixture seen in almost all populations and loses information.

The problem of interpreting log ratio composition association results has been recognized and different approaches suggested. Linear algebra ‘purists’ suggest that since these transformed bases are optimal for the problem, we should reorient our thinking to these variables; the contrarian view is that the mathematical structures are deficient if they cannot enable estimation or inference against physically meaningful hypotheses. Constrained to these bounds, tests of the association of an outcome under conditions of part-wise isoancestry is not possible. Genome compositions do not have an obvious physical logarithmic dependency between the admixed segments of different ancestries during a zygote conception event. To surmount these difficulties, we implemented a new method based on average marginal effects to test genetic admixing isoancestry hypotheses that are not readily amenable under a log ratio basis set of genetic ancestry. This couples an optimal ilr basis set for model fitting, with an epidemiology technique of deriving post-estimation average marginal effects^60^ which averages over the observed covariate distribution.

While the statistical evidence was strong for association of log ratios and isoancestry changes in genetic composition with the MPO outcome, the variance explained by genetic ancestry in the MPO trait was quite small at a few percent, but the greatest variance explained was found in the REC-Ca (4%) and MCC (3%) study groups with likely higher enteropathogen load at the time of stool sampling. This is consistent with eQTL studies of immune response to infection which have suggested that some proportion of ‘missing’ eQTLs are likely context-dependent ^61–64^. Here, the necessary context is enteric infection and suggests that fecal MPO levels may better reflect genetic influences under conditions of active infection. For other diseases that may employ MPO as a fecal biomarker, this is an important consideration; the response may be more important than ambient levels. Directly mirroring our results, it has been shown that the apparent heritability of an infectious disease phenotype depends heavily on the specific disease stage ^65^. The other point to emphasize is that we have not measured absolute heritability in this study; the association with genetic ancestry attempts to attribute differences in heritability between continental populations.

In our study, fecal MPO indirectly measured enteric neutrophil activity which is controlled by gut-specific innate immune responses. The association of increased MPO with increased relative African ancestry is particularly interesting in that African ancestry is known to be associated with persistent reduced white blood cell count, primarily driven by neutrophils (benign ethnic neutropenia) ^66^. This was later shown to be likely driven by differences in ancestral allele frequencies at the Duffy Null polymorphism (rs2814778) ^67^.

Our analyses were repeated with principal component covariates to adjust for SES to establish whether the ancestral differences are due to SES stratification, likely as a consequence of racism. We found a similar result to non-SES adjusted data, albeit with a much smaller sample size in only two studies. The association of relative AFR ancestry with log(MPO) was attenuated but still significant. Caution is necessary since race and genetic ancestry are highly confounded and extremely complex to disentangle. Our goal here was not to perform a sociologically rigorous analysis but to establish the plausibility of a genetic association independent of these other factors.

The major limitation in this study, beyond sample size, was the cross-sectional stool sampling in three of the study groups that will not have captured the time-dependent and dynamic nature of the excreted biomarker. The single longitudinal study also collected stools sample to a protocol surveillance schedule rather than responding to an incipient enteric infection and diarrheal event hence we did not measure acute MPO response to an infection. Field collection of stool samples and accompanying event data is time-consuming and expensive and is one reason why fecal biomarkers have been under-studied.

In summary we have shown that host genetics is a possible unrealized explanatory factor for the variation in fecal MPO biomarker levels. This has application in use of MPO as a biomarker for pediatric enteric infectious disease but also in other diseases such as inflammatory bowel disease and other colitides.

## Declarations

### Ethics Statement

Study protocols were reviewed and approved by the Institutional Review Board for Health Sciences Research of the University of Virginia; and the Research Ethics Committee of Universidade Federal de Cearà.

### Data Availability Statement

The datasets generated and/or analyzed during the current study are available in the NIH dbGaP repository, contiguous accession numbers phs003172.v1.p1 through to phs003175.v1.p1, with web links http://www.ncbi.nlm.nih.gov/projects/gap/cgi-bin/study.cgi?study_id=phs003172.v1.p1 (and similar).

### Availability of data and materials

The datasets generated and/or analyzed during the current study are available in the dbGaP repository, contiguous accession numbers phs003172.v1.p1 through to phs003175.v1.p1, with web links http://www.ncbi.nlm.nih.gov/projects/gap/cgi-bin/study.cgi?study_id=phs003172.v1.p1 (and similar).

### Authors’ contributions

JCM, RLG, AAML conceived and/or designed the study. AH, UN, AAML acquired data. JCM, AH, UN analyzed data. JCM wrote the paper and all authors reviewed the manuscript.

## Acknowledgements

We very gratefully acknowledge the participation of all study families for their support of this research.

